# Hospitalization of mild cases of community-acquired pneumonia decreased more than severe ones during the COVID-19 epidemic

**DOI:** 10.1101/2021.03.18.21253861

**Authors:** Hiroyuki Nagano, Daisuke Takada, Jung-ho Shin, Tetsuji Morishita, Susumu Kunisawa, Yuichi Imanaka

## Abstract

**Objective:** The epidemic of the coronavirus disease 2019 (COVID-19) has affected the entire health care systems. Our aim was to assess the impact of the COVID-19 epidemic on the number and severity of cases for community-acquired pneumonia (CAP) in Japan.

**Methods:** Using claims data from the Quality Indicator/Improvement Project (QIP) database, we included urgent cases of inpatients for CAP from August 1, 2018, to July 30, 2020. We compared the monthly ratio of inpatient cases from August 2018 to July 2019 and August 2019 to July 2020 as a year-over-year comparison. We also compared this ratio according to the severity score “A-DROP” and performed an interrupted time series analysis (ITS) to evaluate the impact of the COVID-19 epidemic on the monthly number of inpatient cases.

**Results:** A total of 67,900 inpatient cases for CAP in 262 hospitals were included. During the COVID-19 epidemic (defined as the period between March and July 2020), the number of inpatient cases for CAP drastically decreased during the epidemic compared with the same period in the past year (−48.1%), despite only a temporary reduction in the number of other urgent admissions. The number of inpatient cases decreased according to the severity of pneumonia. Milder cases showed a greater decrease in the year-over-year ratio than severe ones (mild −55.2%, moderate −45.8%, severe −39.4%, and extremely severe − 33.2%). The ITS analysis showed that the COVID-19 epidemic reduced the monthly number of inpatient cases for CAP significantly (estimated decrease: −1233 cases; 95% CI, −521 to −1955).

**Conclusions:** Our study showed a significant reduction in the number of inpatient cases for CAP during the COVID-19 epidemic in Japan. The milder cases showed a greater decrease in the year-over-year ratio of the number of inpatient cases.

## Introduction

The coronavirus disease 2019 (COVID-19) was first recognized in early December 2019 in Wuhan, China and spread globally in 2020 (Zhu et al., 2020). In Japan, the government focused on controlling the clusters of infected cases by preventing the spread in closed spaces, crowded areas, and close-contact settings (Shimizu et al., 2020). However, the daily number of new COVID-19 infections increased in March 2020. The Japanese government declared, then, a state of emergency and requested self-quarantine and social distancing in seven prefectures on April 7 and 47 prefectures on April 16 (Prime Minister of Japan and His Cabinet, 2020). Consequently, the COVID-19 epidemic has changed the lifestyle of many Japanese as individual hygiene, maintenance of social distance, and suspension of large-scale gatherings were strongly encouraged.

Community-acquired pneumonia (CAP) is defined as a lower respiratory tract infection that is acquired outside of hospital setting (Metlay et al., 2019). CAP is one of the most common infectious disease responsible for hospital admission and mortality, especially in the elderly. With the COVID-19 epidemic, the decrease of inpatient and outpatient cases for CAP were reported (Wu et al., 2020; Yamamoto et al., 2020). However, only a few studies focused on finding the type of CAP patients who were impacted by the COVID-19 epidemic. Therefore, we performed a retrospective cohort study using a large-scale Japanese database. Our study sought to evaluate the impact of the COVID-19 epidemic on the number and severity of inpatients for CAP in Japan.

## Methods

### Data source

For this retrospective cohort study, we extracted Diagnosis Procedure Combination (DPC) data from the Quality Indicator/Improvement Project (QIP) database. The QIP is administered by the Department of Healthcare Economics and Quality Management in Kyoto University (Hamada et al., 2012), which regularly collects DPC data from acute care hospitals in Japan that voluntarily participate in the project. These hospitals include both public and private facilities of various sizes. In 2019, the number of general beds at the participating hospitals ranged from 30 to 1,151 (excluding psychiatric, infectious diseases, and tuberculosis beds according to the Japanese classification of hospital beds). The DPC/per-diem payment system (PDPS) is a Japanese prospective payment system applied to acute care hospitals. In 2018, a total of 1,730 hospitals used the DPC/PDPS, accounting for 54% (482,618 out of 891,872) of all the general hospital beds in Japanese hospitals (Ministry of Health, Labour and Welfare, 2018; Ministry of Health, Labour and Welfare, 2020). The DPC consists of several data files, including forms 1, 3, and 4 and files D, E, F, H, and K (Ministry of Health, Labour and Welfare, 2020). For this study, we used form 1, file E, and file F. Form 1 contains discharge summaries, which include the International Classification of Diseases 10^th^ Revision (ICD-10) codes classifying main diagnosis, the trigger diagnosis, the most and second-most medical-resource-intensive diagnoses, up to 10 comorbidities, and 10 complications during hospitalization. Form 1 also includes the following patient details: age, sex, body mass index (BMI), components of the Barthel index (independence of feeding, bathing, grooming, dressing, bowels, bladder, toilet use, transfers from bed to chair, mobility on level surfaces, and stairs), medical procedures, the pneumonia severity score according to the A-DROP scoring system (a modified version of the CURB-65 scoring in Japan) at the onset, and the type of pneumonia (community-acquired pneumonia, nosocomial pneumonia, or diagnosis other than pneumonia). Files E and F contain information on all medical services, medications, and equipment for both inpatients and outpatients. File E and F include the name of the diagnosis, the start date of diagnosis, and the date of visit for the outpatients.

### Study population

This study includes cases of patients aged 18 years and older and who were hospitalized urgently for pneumonia between August 1^st^, 2018 and July 31^st^, 2020. We defined urgent admission as unplanned hospital admission and pneumonia as stipulated in the ICD-10 code in the 2013 version: J10.0, J11.0, J12.x-J18.x, A48.1, B01.2, B05.2, B37.1, and B59.x. The exact contents of the ICD-10 codes are specified in table S1 in the supplemental data. CAP was identified using the corresponding variable in the DPC data. We included inpatient cases with ICD-10 codes in the following data fields: main diagnosis, trigger diagnosis, and most medical resource-intensive diagnosis. We extracted the urgent admissions for diagnosis other than pneumonia (other urgent admissions) for comparison. We also extracted the outpatient CAP cases newly diagnosed with pneumonia from file E and F in the same period to evaluate the trend of overall patients with CAP. Newly diagnosis of pneumonia was defined as the coincidence between the start date of diagnosis and the date of visit.

### Year-over-year comparison

The primary outcome of interest was the change in the monthly number of inpatient CAP cases during the COVID-19 epidemic (defined as between March and July 2020). We retrospectively compared the monthly ratio of inpatient cases between August 2018 to July 2019 and August 2019 to July 2020 as a year-over-year comparison. The year-over-year comparisons were also performed for other urgent admissions and outpatient pneumonia cases.

We also performed the year-over-year comparison of the inpatient cases in each group of the A-DROP scoring system from March to July in 2019 and 2020. The A-DROP system is a 6-point scale (0–5) that assess the clinical severity of CAP according to the following parameters: (i) age (male ≥ 70 years, female ≥ 75 years), (ii) dehydration (blood urea nitrogen (BUN) ≥ 210 mg/L), (iii) respiratory failure (arterial oxygen saturation (SpO2) ≤ 90% or partial pressure of oxygen in arterial blood (PaO2) ≤ 60 mmHg), (iv) orientation disturbance (confusion), and (v) low blood pressure (systolic blood pressure (SBP) ≤ 90 mmHg) (Shindo et al., 2008). We divided the A-DROP score into four severity classes (mild, 0; moderate, 1-2; severe, 3; and extremely severe, 4-5). Specifically, we used the locally estimated scatterplot smoothing (LOESS) to visualize smooth trend curves.

### Descriptive analysis

We compared the characteristic of inpatient cases from March to July 2019 and 2020. The characteristics were described for the overall cases and each group of the A-DROP scoring system. We presented the body mass index, the Carlson Comorbidity index, and the Barthel index variables because they are associated with the mortality of inpatient pneumonia cases (Nguyen et al., 2019; Murcia et al., 2010; Takada et al., 2020). Age and hospital length of stay were expressed as median values (interquartile range). We compared these variables between the two groups using Mann-Whitney U tests. The categorical data (such as sex) were compared using chi-square or Fisher’s exact tests. All statistical analyses were performed with R version 3.6.0 (R Foundation for Statistical Computing, Vienna, Austria).

### Interrupted time series analyses for changes in case numbers

An interrupted time series analysis (ITS) was performed to evaluate the impact of the COVID-19 epidemic on the monthly number of inpatient cases (Bernal et al., 2017). We statistically tested the changes in the number of monthly cases after adjusting for seasonality using a Fourier term. We hypothesized that the COVID-19 epidemic would reduce the monthly number of cases after March 2020 since this was when the first wave of COVID-19 started (Shimizu et al., 2020). We also conducted a sensitivity analysis. To assess the sensitivity of our findings to a change in time point, we changed this parameter. The candidate intervention time points were as follows: 1) February 2020, in Japan, the Ministry of Health, Labour and Welfare of Japan requested pharmacies and supermarkets to impose restrictions on purchases for the shortage of face masks and disinfectants (Sakamoto et al., 2020), and 2) April 2020, the Government of Japan declared the state of emergency (Prime Minister of Japan and His Cabinet, 2020). An interrupted time series analysis was also performed to evaluate the impact of the COVID-19 epidemic on the monthly number of outpatient cases.

## Results

We included a total of 67,900 inpatient CAP cases in 262 hospitals. We also extracted 1,425,133 other urgent admission and 48,288 outpatient CAP cases. The monthly numbers of cases and year-over-year ratios are displayed in table S2 in the supplemental material. During the COVID-19 epidemic, the number of cases decreased compared with the same period the year before (inpatient CAP cases −48.1%, other urgent admissions −9.1%, and outpatient CAP cases −26.7%). The year-over-year ratios of inpatient CAP cases and other urgent admissions are illustrated in Figure 1. During the COVID-19 epidemic, the number of inpatient CAP cases drastically decreased despite only a temporary reduction of the number of other urgent admissions.

**Figure 1:**
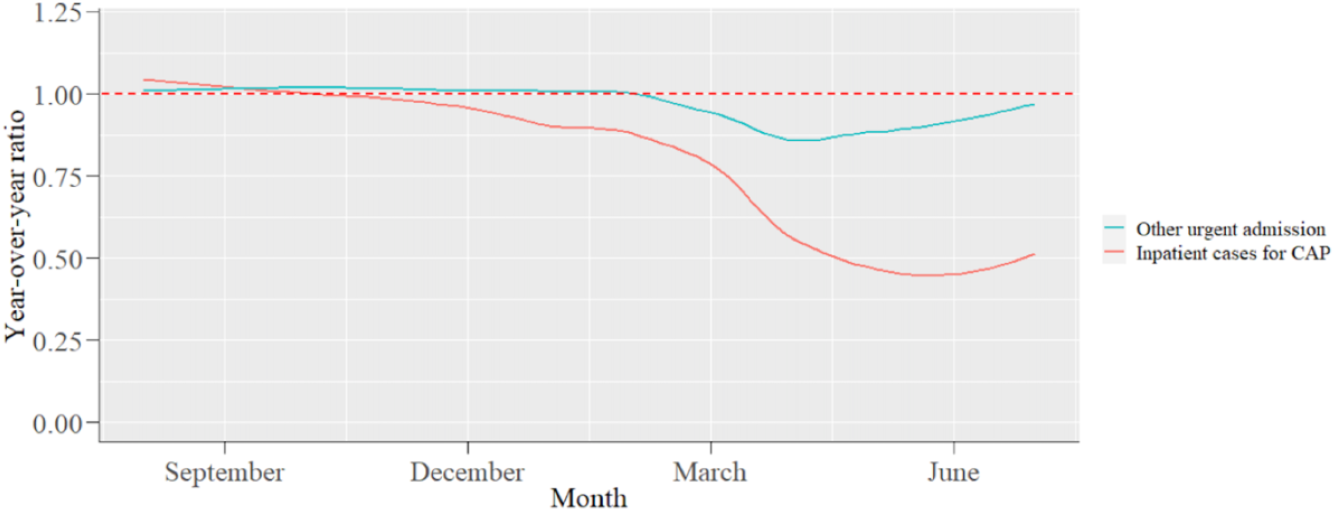
**Year-over-year ratio of the number of other urgent admission for diagnosis other than pneumonia and inpatient cases for CAP.** Other urgent admission: urgent admission for diagnosis other than pneumonia. CAP: Community-acquired pneumonia.

Figure 2 shows the year-over-year ratios of the inpatient cases in each group of the A-DROP scoring system. The monthly numbers of cases and year-over-year ratios in each group of the A-DROP scoring system are displayed in table S3 in the supplemental material. During the COVID-19 epidemic, the year-over-year ratios for the milder inpatient cases decreased more than for the severe cases (mild −55.2%, moderate −45.8%, severe −39.4%, extremely severe −33.2%).

**Figure 2:**
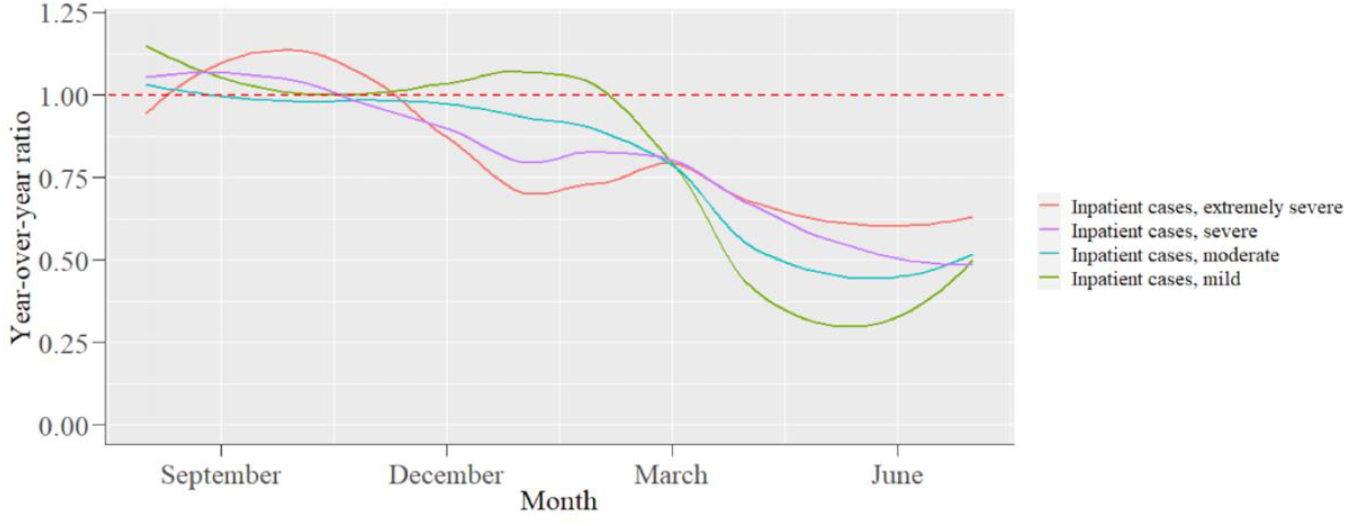
**Year-over-year ratio of the number of inpatient cases for community-acquired pneumonia in each group of A-DROP scoring system**

Table 1 displays the baseline characteristics of the inpatient cases in each group. Especially, the ratio of inpatient cases with chronic pulmonary disease in the mild group was lower than during the same period in 2019 (21.5% vs 17.5%). For overall cases, the in-hospital mortality was more elevated during the COVID-19 epidemic than the same period in 2019 (6.4% vs 9.3%). The ratio of several categories associated with pneumonia mortality, such as the Charlson Comorbidity index≧1 (71.6% to 73.8%), the Charlson Comorbidity index≧2 (40.8% to 43.9%), the Barthel index≦80 (50.7% to 54.2%), and BMI<17kg/m2 (14.1% to 16.7%), was also increased. In each group, the ratio of in-hospital death and these categories were also increased.

**Table 1:**
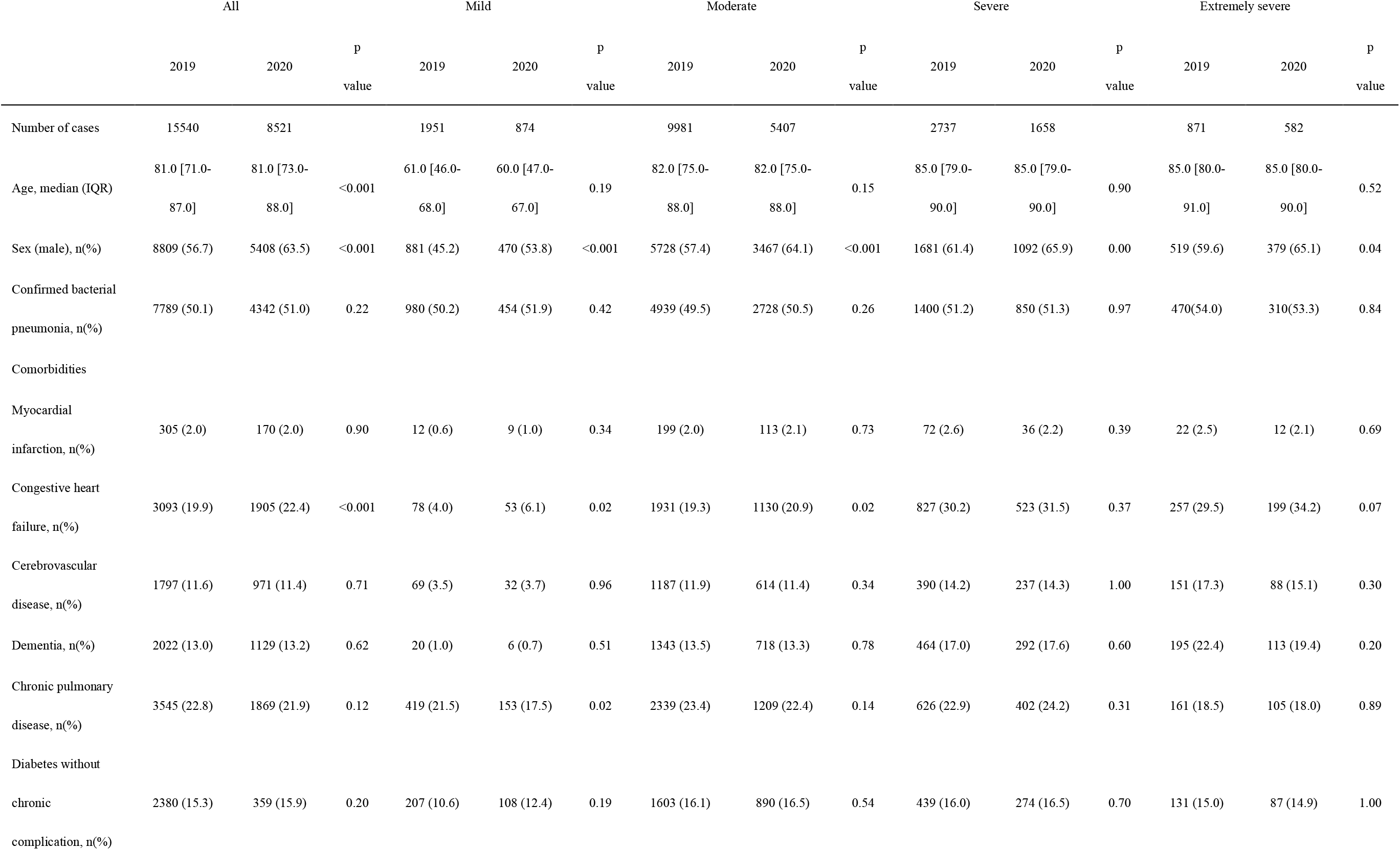
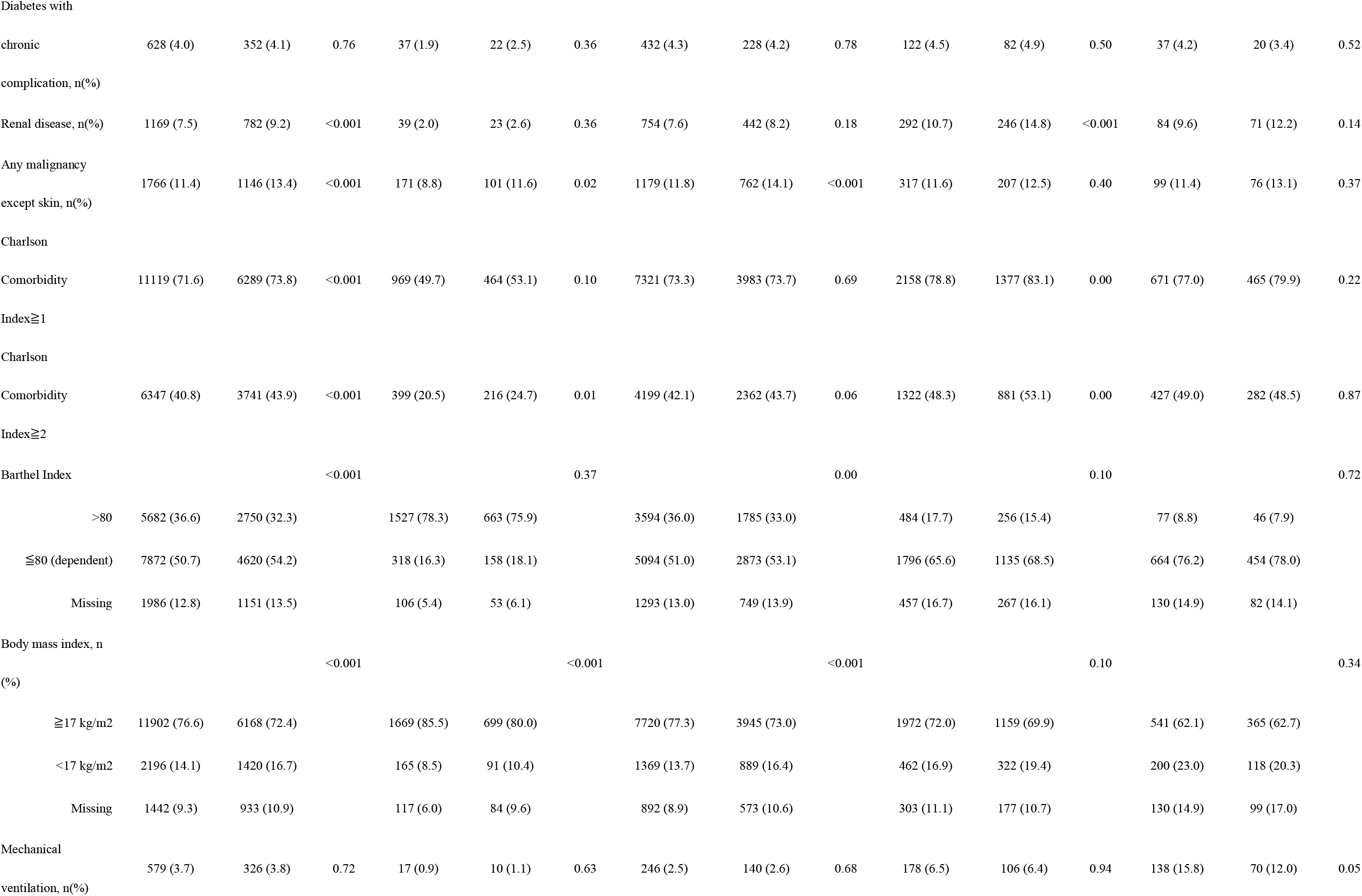
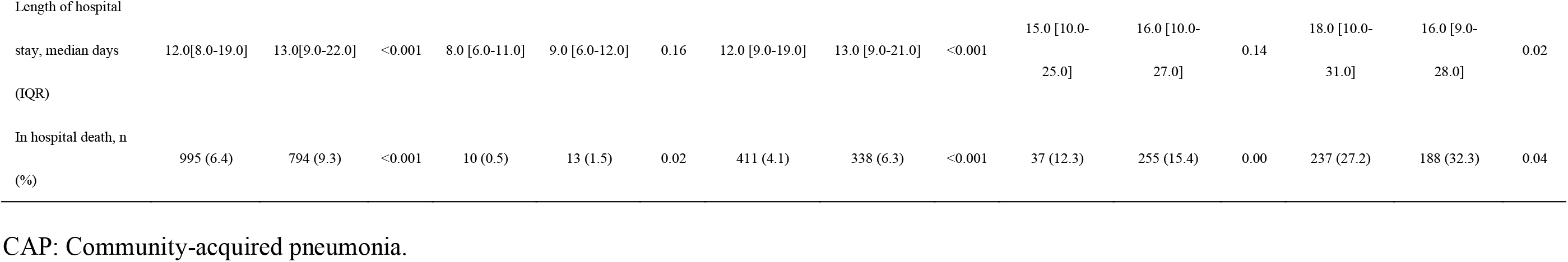
The characteristics of inpatient cases for CAP in each group of the A-DROP scoring system between March and July in 2019 and 2020.

The ITS analysis showed a significant reduction of the monthly number of inpatient CAP cases (estimated decrease: −1233 cases; 95% CI, −521 to −1955) and outpatient CAP cases (estimated decrease: −1808 cases; 95% CI, −656 to −2960). Our findings were largely unaffected by the change of the time point as shown in Table S4 in the supplemental material.

## Discussion

In this study, we used a large-scale administrative database to analyze the impact of the COVID-19 epidemic on the number and severity of inpatient CAP cases. Our main findings were as follows: first, a significant reduction of the number of inpatient CAP cases despite an only temporary reduction of the number of other urgent admissions during the COVID-19 epidemic; second, a greater decrease of the year-over-year ratio in milder cases; third, a significant decrease of the number of inpatient cases and outpatient CAP cases.

The decrease of inpatient CAP cases was consistent with previous reports (Wu et al., 2020; Yamamoto et al., 2020). Our study revealed that milder cases of pneumonia were associated with a greater decrease in the year-over-year ratio of the number of inpatient cases. Many patients were reluctant to seek medical evaluation for fear of exposure to COVID-19 in the hospital in other departments, including pediatrics and cardiology (Hammad et al., 2020; Lazzerini et al., 2020). The Japanese government urged patients with light symptoms to stay at home on February 22, 2020 (Sakamoto et al., 2020). Therefore, the patients with mild symptoms might have the same tendency to avoid medical visits in our study population.

Moreover, the change of lifestyle itself might reduce the onset of pneumonia. Our study included inpatient cases in the elderly with a median age of 80. The main pathogens responsible for pneumonia in the elderly include *Streptococcus pneumonia, Haemophilus influenzae, and Enterobacteriaceae* (Janssens, 2005). The aspiration of microorganisms inside the oral cavity is also a major cause of pneumonia in the elderly because the elderly have risk factors including impaired swallowing and cough reflex (Mandell and Niederman, 2019). These bacteria do not spread directly from human to human. The COVID-19 epidemic changed the patients’ lifestyle by encouraging individual-level hygiene (wearing a mask outside, keeping social distance, disinfecting with alcohol, and washing hands frequently) and community-level prevention measures (promotion of remote work, suspension of mass gathering, and school closure) (Abe et al., 2021). Limiting family visits were carried out in many aged care facilities in Japan (ABC News, 2020). These lifestyle changes might prevent many person-to-person disease transmissions, such as respiratory viral diseases (Sakamoto et al., 2020; de Souza Luna et al., 2020; Sunagawa et al., 2021; Jones, 2020) and tuberculosis (Komiya et al., 2020). A respiratory viral infection is an important risk factor for bacterial pneumonia because respiratory viruses affect the pulmonary defense of the host and impair bacterial clearance (Prasso and Deng, 2017). A previous report showed a significant increase in hospitalizations due to pneumococcal pneumonia during the 2009 influenza pandemic (Weinberger et al., 2012). The behavior change due to the COVID-19 infection control may prevent viral respiratory infection and consequently bacterial pneumonia. Therefore, it might contribute to the decrease in the onset of pneumonia during the COVID-19 epidemic. Our study also showed the ratio of inpatient cases with milder chronic pulmonary disease decreased during the COVID-19 epidemic compared with the same period in 2019. Previous studies revealed that the change in patient behavior and physical environment during the COVID-19 epidemic could contribute to the decrease in hospital admissions for acute exacerbation of chronic obstructive pulmonary diseases and asthma (Abe et al., 2021; Chan et al., 2020). These changes could also contribute to the decrease in inpatient CAP cases, especially for the patients with milder diseases. Other possibilities included the shortage of hospital beds and manpower restricted to hospitalization (Zhou et al., 2020). However, table S2 revealed that the number of other urgent admissions recovered to about 90%. This result suggests the absence of a large number of restrictions. Additionally, the decline in outpatient CAP cases cannot be explained by the restrictions in hospitalization.

Our study revealed that for overall cases and each group of A-DROP scoring system, the in-hospital mortality was more elevated during the COVID-19 epidemic than the same period in 2019. A higher Charlson Comorbidity index, functional dependence (Barthel index≦80 points), and BMI<17kg/m2 were associated with higher mortality in patients with pneumonia (Nguyen et al., 2019; Murcia et al., 2010; Takada et al., 2020). As shown in Table 1, the ratios in these categories increased during the COVID-19 epidemic. These changes might contribute to the elevated mortality in each group of the A-DROP scoring system during the COVID-19 epidemic.

Our study has several limitations. First, the study population was restricted to patients in hospitals that voluntarily participated in the QIP. Therefore, a selection bias cannot be excluded. Second, the diagnoses of the outpatient pneumonia cases recorded in files E and F were not classified as in form 1 and may not be sufficiently robust. However, files E and F contained the start date of diagnosis and date of visit and allowed us to extract the outpatient cases newly diagnosed with pneumonia. Therefore, the trend for the number of outpatient pneumonia cases was approximately assessed. Despite these limitations, our research provided important information on the impact of the COVID-19 epidemic on CAP inpatients using a large-scale Japanese database. Further studies are warranted to identify the long-term impact of the decrease of inpatient CAP cases on patient outcomes and health care systems.

In conclusion, we showed a marked reduction of inpatient CAP cases during the COVID-19 epidemic in Japan using large-scale administrative data. The decrease in the year-over-year ratio of the number of inpatient cases was greater for the milder pneumonia cases.

## Data Availability

Data are available upon reasonable request. Please contact the corresponding author.

## Funding

This study was supported by JSPS KAKENHI Grant Numbers JP19H01075 from the Japan Society for the Promotion of Science (https://www.jsps.go.jp/english/e-grants/), Health Labour Sciences Research Grants from the Ministry of Health, Labour and Welfare, Japan, Grant Numbers 20HA2003 (https://www.mhlw.go.jp/stf/seisakunitsuite/bunya/hokabunya/kenkyujigyou/index.html), and by GAP Fund Program of Kyoto University, GAP Fund Program Type B (http://www.venture.saci.kyoto-u.ac.jp/?page_id=83#gp) to Y. I.

The funders played no role in the study design, data collection and analysis, decision to publish, or preparation of the manuscript.

## Ethical considerations

This reasearch was conducted according to the Ethical Guidelines for Medical and Health Research Involving Human Subjects of the MHLW, Japan (a provisional translation is available from: https://www.mhlw.go.jp/file/06-Seisakujouhou-10600000-Daijinkanboukouseikagakuka/0000080278.pdf). In the accordance of the Guidelines, informed consent was not required for research not utilizing human biological specimens and information utilized in the research has been anonymized. The Ethics Committee, Graduate School of Medicine, Kyoto University approved the study (approval number: R0135).

## Potential conflicts of interest

The authors: No reported conflicts of interest.

## Contribution

HN: Conceptualization, methodology, software, formal analysis, writing - original draft, writing - review & editing, visualization; DT: Conceptualization, methodology, software, validation, investigation, writing - review & editing; JS: Conceptualization, software, validation, investigation, data curation, writing - review & editing; TM: Conceptualization, methodology, software, validation, investigation, writing - review & editing; SK: Conceptualization, validation, investigation, data curation, resources, writing - review & editing; YI: Conceptualization, validation, investigation, resources, writing - review & editing, supervision, project administration, funding acquisition

## Acknowledgement

We thanked the staff of participating hospitals in QIP.

## Declaration of interests

The authors declare that they have no known competing financial interests or personal relationships that could have appeared to influence the work reported in this paper.

## Notes

### Competing Interest Statement

The authors have declared no competing interest.

### Author Declarations

The Ethics Committee, Graduate School of Medicine, Kyoto University approved the study (approval number: R0135).

