## Supplementary material for "Hospitalization of mild cases of community-acquired pneumonia decreased more than severe ones during the COVID-19 epidemic": Table S1

**Table S1: ICD-10 code into each category.**

| ICD-10 code | Disease |
|---|---|
| J10.0, J11.0 | Influenza with pneumonia |
| J12.x | Viral pneumonia |
| J13 | Pneumococcal pneumonia |
| J14 | Pneumonia due to haemophilus influenzae |
| J15.x | Pneumonia due to other specified bacteria |
| J16.x | Pneumonia due to other infectious pathogens, not elsewhere classified |
| J17.x | Pneumonia due to diseases classified elsewhere |
| J18.x | Pneumonia, unspecified organism |
| A48.1 | Legionellosis |
| B01.2 | Varicella pneumonia |
| B05.2 | Measles, complicated with pneumonia |
| B37.1 | Pulmonary candidiasis |
| B59 | Pneumocystosis |
