## Supplementary material for "Hospitalization of mild cases of community-acquired pneumonia decreased more than severe ones during the COVID-19 epidemic": Table S2

**Table S2: The number of cases per month and year-over-year ratio for inpatient cases for CAP, other urgent admission and outpatient cases for CAP.**

| **Inpatient cases for CAP** | August | September | October | November | December | January | February | March | April | May | June | July | Total |
|---|---|---|---|---|---|---|---|---|---|---|---|---|---|
| 2018/8-2019/7 | 3002 | 2539 | 2940 | 3027 | 3222 | 4692 | 2972 | 2865 | 3186 | 3517 | 2850 | 3122 | 37934 |
| 2019/8-2020/7 | 3100 | 2703 | 2758 | 3005 | 3227 | 3915 | 2737 | 2303 | 1738 | 1490 | 1449 | 1541 | 29966 |
| year-over-year ratio | 1.03 | 1.06 | 0.94 | 0.99 | 1.00 | 0.83 | 0.92 | 0.80 | 0.55 | 0.42 | 0.51 | 0.49 | 0.79 |
| **Other urgent admission** | August | September | October | November | December | January | February | March | April | May | June | July | Total |
| 2018/8-2019/7 | 63697 | 57370 | 61498 | 59671 | 61665 | 63729 | 54132 | 59124 | 60362 | 61127 | 58062 | 62244 | 722681 |
| 2019/8-2020/7 | 63875 | 59117 | 61828 | 60393 | 63402 | 62267 | 55593 | 56109 | 50800 | 53361 | 57212 | 58495 | 702452 |
| year-over-year ratio | 1.00 | 1.03 | 1.01 | 1.01 | 1.03 | 0.98 | 1.03 | 0.95 | 0.84 | 0.87 | 0.99 | 0.94 | 0.97 |
| **Outpatient cases for CAP** | August | September | October | November | December | January | February | March | April | May | June | July | Total |
| 2018/8-2019/7 | 5236 | 4548 | 5357 | 5197 | 5384 | 7000 | 4901 | 5058 | 5706 | 6346 | 5327 | 5642 | 65702 |
| 2019/8-2020/7 | 5341 | 4951 | 5073 | 5201 | 5647 | 6358 | 5182 | 5486 | 5148 | 3947 | 3757 | 4020 | 60111 |
| year-over-year ratio | 1.02 | 1.09 | 0.95 | 1.00 | 1.05 | 0.91 | 1.06 | 1.08 | 0.90 | 0.62 | 0.71 | 0.71 | 0.91 |

Other urgent admission: urgent admission for diagnosis other than pneumonia.

CAP: Community acquired pneumonia.
