## Supplementary material for "Hospitalization of mild cases of community-acquired pneumonia decreased more than severe ones during the COVID-19 epidemic": Table S3

Table S3: **The number of cases per month and year-over-year ratio for inpatient cases for CAP according to the A-DROP scoring system.**

| **Mild** | August | September | October | November | December | January | February | March | April | May | June | July | Total |
|---|---|---|---|---|---|---|---|---|---|---|---|---|---|
| 2018/8-2019/7 | 368 | 293 | 395 | 399 | 390 | 471 | 306 | 332 | 421 | 457 | 371 | 370 | 4573 |
| 2019/8-2020/7 | 418 | 323 | 370 | 398 | 428 | 454 | 347 | 254 | 174 | 127 | 142 | 177 | 3612 |
| year-over-year | 1.14 | 1.10 | 0.94 | 1.00 | 1.10 | 0.96 | 1.13 | 0.77 | 0.41 | 0.28 | 0.38 | 0.48 | 0.79 |
| **Moderate** | August | September | October | November | December | January | February | March | April | May | June | July | Total |
| 2018/8-2019/7 | 1976 | 1699 | 1880 | 1943 | 2010 | 2839 | 1838 | 1827 | 2018 | 2296 | 1833 | 2007 | 24166 |
| 2019/8-2020/7 | 2024 | 1750 | 1734 | 1890 | 2088 | 2393 | 1758 | 1436 | 1084 | 958 | 941 | 988 | 19044 |
| year-over-year | 1.02 | 1.03 | 0.92 | 0.97 | 1.04 | 0.84 | 0.96 | 0.79 | 0.54 | 0.42 | 0.51 | 0.49 | 0.79 |
| **Severe** | August | September | October | November | December | January | February | March | April | May | June | July | Total |
| 2018/8-2019/7 | 492 | 409 | 522 | 525 | 603 | 987 | 595 | 531 | 556 | 571 | 499 | 580 | 6870 |
| 2019/8-2020/7 | 504 | 474 | 490 | 551 | 515 | 797 | 474 | 457 | 357 | 303 | 264 | 277 | 5463 |
| year-over-year | 1.02 | 1.16 | 0.94 | 1.05 | 0.85 | 0.81 | 0.80 | 0.86 | 0.64 | 0.53 | 0.53 | 0.48 | 0.80 |
| **Extremely severe** | August | September | October | November | December | January | February | March | April | May | June | July | Total |
| 2018/8-2019/7 | 166 | 138 | 143 | 160 | 219 | 395 | 233 | 175 | 191 | 193 | 147 | 165 | 2325 |
| 2019/8-2020/7 | 154 | 156 | 164 | 166 | 196 | 271 | 158 | 156 | 123 | 102 | 102 | 99 | 1847 |
| year-over-year | 0.93 | 1.13 | 1.15 | 1.04 | 0.89 | 0.69 | 0.68 | 0.89 | 0.64 | 0.53 | 0.69 | 0.60 | 0.79 |

CAP: Community acquired pneumonia.
