## Supplementary material for "Hospitalization of mild cases of community-acquired pneumonia decreased more than severe ones during the COVID-19 epidemic": Table S4

**Table S4: Interrupted time series analysis for in patient cases and outpatient cases for CAP.**

| | Time point | Estimate | Lower 95%CI | Upper 95% CI |
|---|---|---|---|---|
| Inpatient case for CAP | February 2020 | -1337 | -1991 | -684 |
| | March 2020 | -1233 | -1955 | -521 |
| | April 2020 | -1309 | -2047 | -571 |
| Outpatient cases for CAP | February 2020 | -1827 | -2951 | -703 |
| | March 2020 | -1808 | -2960 | -656 |
| | April 2020 | -2292 | -3329 | -1256 |

CAP: Community acquired pneumonia.

Estimate: estimated decrease in the number of cases in the interrupted time series analysis.
